# The mobility gap: estimating mobility levels required to control Canada’s winter COVID-19 surge

**DOI:** 10.1101/2021.01.28.21250622

**Authors:** Kevin A. Brown, Jean-Paul R. Soucy, Sarah A. Buchan, Shelby L. Sturrock, Isha Berry, Nathan M. Stall, Peter Jüni, Amir Ghasemi, Nicholas Gibb, Derek R. MacFadden, Nick Daneman

## Abstract

**Background:** Non-pharmaceutical interventions remain a primary means of suppressing COVID-19 until vaccination coverage is sufficient to achieve herd immunity. We used anonymized smartphone mobility measures in seven Canadian provinces to quantify the mobility level needed to suppress COVID-19 (mobility *threshold*), and the difference relative to current mobility levels (mobility *gap*).

**Methods:** We conducted a longitudinal study of weekly COVID-19 incidence from March 15, 2020 to January 16, 2021, among provinces with 20 COVID-19 cases in at least 10 weeks. The outcome was weekly growth rate defined as the ratio of current cases compared to the previous week. We examined the effects of average time spent outside the home (non-residential mobility) in the prior three weeks using a lognormal regression model accounting for province, season, and mean temperature. We calculated the COVID-19 mobility threshold and gap.

**Results:** Across the 44-week study period, a total of 704,294 persons were infected with COVID-19. Non-residential mobility dropped rapidly in the spring and reached a median of 36% (IQR: 31,40) in April 2020. After adjustment, each 5% increase in non-residential mobility was associated with a 9% increase in the COVID-19 weekly growth rate (ratio=1.09, 95%CI: 1.07,1.12). The mobility gap increased through the fall months, which was associated with increasing case growth.

**Interpretation:** Mobility strongly and consistently predicts weekly case growth, and low levels of mobility are needed to control COVID-19 through winter 2021. Mobility measures from anonymized smartphone data can be used to guide the provincial and regional implementation and loosening of physical distancing measures.

## Background

The global toll of Coronavirus Disease 2019 (COVID-19) continues to grow, despite the promise of recently approved vaccines. A winter surge is occurring in many countries in the Northern Hemisphere, including Canada, that may take a considerable toll before vaccination is sufficiently widespread to achieve herd immunity. Non-pharmaceutical public health interventions (NPIs) including physical distancing remain the primary population-based means of controlling COVID-19 (1). Since early in the second wave, which started in September 2020, polling suggests that most Canadians have supported and adhered to government-directed restrictions (2), and many favor strengthened measures to suppress community transmission of COVID-19 (3).

Severe acute respiratory syndrome coronavirus 2 (SARS-CoV-2), the causative viral agent of COVID-19, is primarily spread through close contact with infected individuals (4). As with any infectious disease, contact rates are a primary driver of COVID-19 transmission (5). Mobility measures capturing human activity through anonymized tracking of smartphones are believed to be reasonable proxies of contact rates outside of one’s own home; these measures can provide more timely and reliable sources of information on contact rates compared to time-use surveys or contact tracing (6–8).

A variety of aggregated, smartphone mobility measures are provided by software developers (e.g., Google and Apple), mobile application developers (e.g., CityMapper), and mobility data vendors (e.g. Veraset, Environics). These measures and data sources have been used to quantify the level and effectiveness of lockdown measures aiming to reduce the spread of COVID-19 (9–11).

Mobility metrics are helpful for gauging the effect of restrictions on behaviour but do not on their own indicate to decision makers if current restrictions are sufficient to curtail the growth of COVID-19. In this study, we examined the association between smartphone mobility measures and growth of COVID-19 in Canadian provinces between March 2020 and January 2021. We sought to quantify the mobility level needed in order to control COVID-19 (mobility *threshold*), as well as the difference between current mobility levels and this threshold (mobility *gap*).

## Methods

### Population and Study Design

We conducted a multi-province longitudinal study of weekly COVID-19 incidence. All province-weeks (since each data point corresponded to a unique week for a given province) with fewer than 20 cases in the prior week were excluded, as small case counts could have been driven by importation rather than local transmission. All provinces or territories with at least 10 weeks with 20 or more cases were eligible for inclusion.

### Outcome

We measured the weekly case counts and test positivity in each province using data from COVID-19 Canada Open Data Working Group (12), in the 44-week period from March 15, 2020 to January 16, 2021. Outcomes were aggregated by week in order to control for day-of-the-week patterns evident in Canadian case reporting data (13). The Open Data Working Group obtains and compiles daily case counts reported across the country by provincial public health agencies, accredited news media, and official social media accounts. Weeks were defined as starting on Sunday and ending Saturday.

Our outcome was the weekly growth ratio measured as the ratio of COVID-19 cases in a given week divided by the number of COVID-19 cases in the prior week. A weekly growth ratio equal to 1 meant that incidence was stable relative to the prior week, weekly growth >1 meant that cases increased. Because surveillance data may be subject to under-detection, we developed a corrected growth ratio, equal to the weekly case growth multiplied by the weekly test positivity growth ratio.

### Mobility Measures

Province-level smartphone mobility data were drawn from open-source Google Community Mobility Reports (14) which are updated daily. Data are collected from select Android users who have turned on the “location history” setting. The primary predictor of interest was the average time spent outside of home (non-residential mobility) in the prior 3 week-period. This lag-period was chosen based on a 10-day buffer around the known peak correlation between mobility and case growth at 11-days (11). We also examined the relative volume of park visits during the same lagged period, as a proportion of non-residential mobility comprise outdoor activities that are low risk for COVID-19 transmission.

For each mobility measure, the baseline level was the median value during the 5-week period from Jan 3–Feb 6, 2020, a period 1-month prior to the first confirmed case of community transmission in Canada (March 5 in British Columbia) and prior to first the first school closures in Canada (March 15 in Ontario). The residential index values were rescaled so that levels in the baseline period represented 100%, with a range from 0% (no non-residential mobility) to values greater than 100% (8,9). Park visit volumes were similarly rescaled. For the purposes of plotting non-residential mobility, we smoothed the index values using a penalized spline with a knot for each 2-week period (15).

### Covariates

In addition to mobility measures, we controlled for week of the year and average temperature in a 3-week lag period (degrees centigrade) of the most populous city of each province, based on Environment Canada data (16).

### Analysis – Regression model

We examined weekly COVID-19 growth using a Gaussian regression model, with the logarithm of the ratio of COVID-19 cases in week T divided by week T-1 as the outcome. Covariate coefficients from this model were exponentiated and represented growth rate ratios (GRR). Factors with GRR values >1 were associated with accelerating growth; factors with GRR values <1 were factors associated with decelerating growth. For the primary and secondary outcome, we developed 3 regression models: (1) a preliminary model that included just non-residential mobility, (2) a base model that added a random intercept for provinces and a penalized spline for the week (with a knot for every 2-month period), and (3) an adjusted model that also accounted for mean temperature in the prior 3 weeks as a linear covariate. Due to high correlation of mean temperature and park mobility (Pearson correlation=0.85), park mobility was examined in a separate analysis. Analogous models were used for the corrected COVID-19 case growth outcome. All models were fit using the mgcv package in R (15).

### Analysis – Mobility Threshold

Using the base model of the association between COVID-19 case growth and mobility (2, above), we estimated the mobility threshold at which COVID-19 case growth would cease to occur. We hypothesized that lower mobility levels may be needed in provinces with larger urban populations in the winter, compared to more rural provinces in the summer (17). This estimated mobility threshold was calculated directly from the base model of case growth by identifying the intercept where the mobility versus COVID-19 case growth association crossed the line of no growth.

### Analysis – Mobility Gap

The mobility gap was defined as the difference between the observed mobility and the mobility threshold. The mobility gap can be interpreted as the estimated incremental reduction in mobility that would have been needed to achieve control COVID-19 case growth in a given province in a given week. The validity of the mobility gap was verified by assessing the association between the estimated mobility gap and actual COVID-19 case growth, across Canada, and for each of the 7 included provinces separately (R^2^).

## Results

### Longitudinal Cohort

Across the 44-week longitudinal study (March 15, 2020 to January 16, 2021), there were 704,294 cases of COVID-19 in Canada. Quebec (N=240,956) and Ontario (N=239,023) had the highest number of COVID-19 cases, while Nova Scotia (N=1,413) and Saskatchewan (N=19,693) had the fewest (Table 1). Quebec, Ontario, Alberta, and British Columbia each had 44 eligible weeks included in the study, while Saskatchewan, Manitoba, and Nova Scotia had 37, 28, and 17 eligible weeks included, respectively.

**Table 1.**
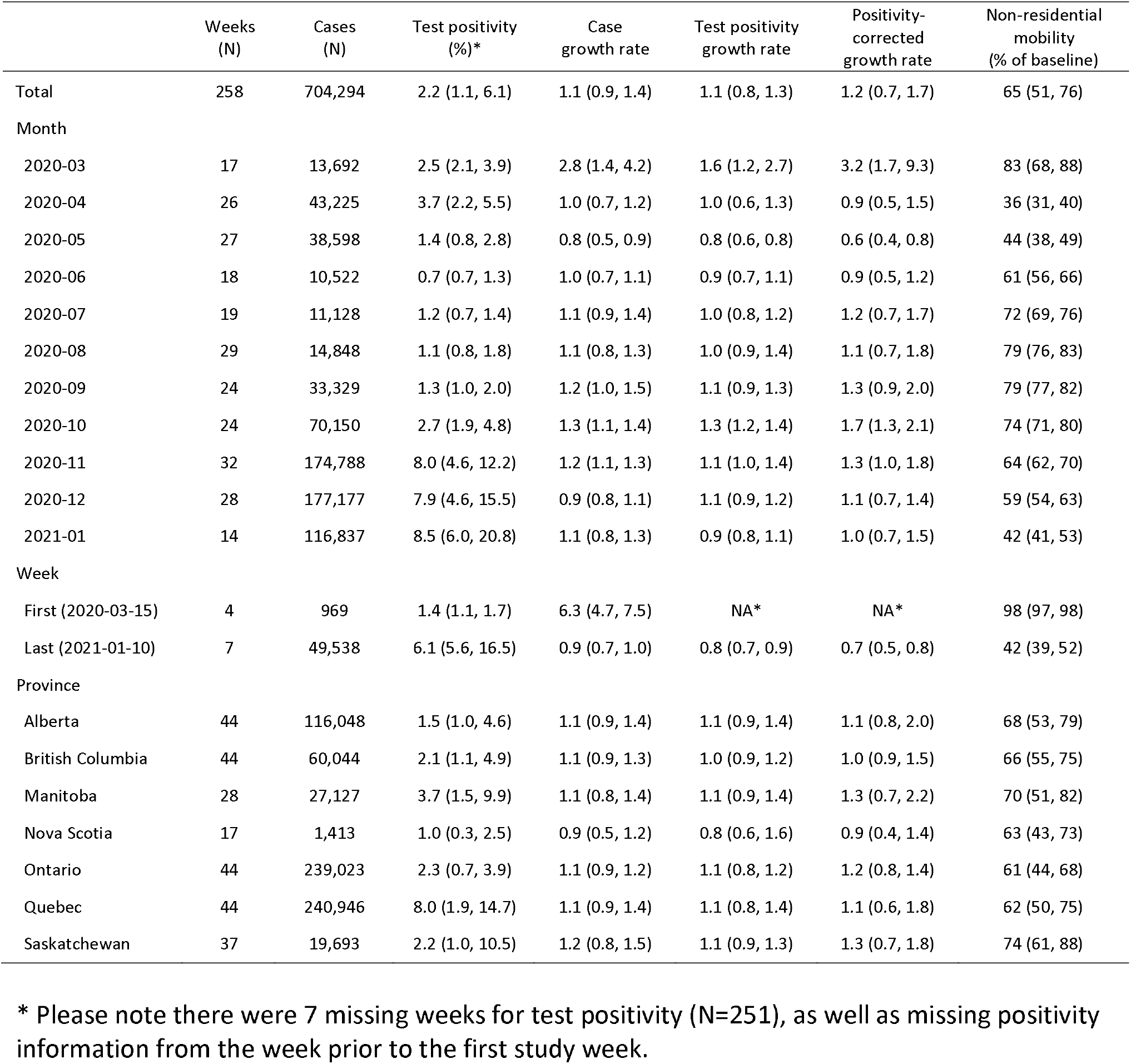
Weekly COVID-19 case growth rates, test positivity, and mobility in Canada, March 15, 2020 to January 16, 2021.

### Mobility

Non-residential mobility dropped rapidly in March to reach a low of 34.3% in the week of April 5^th^, 2020 (median=34.3%, IQR: 29.0% to 35.2%; Figure 1). Mobility increased through the summer of 2020 and reached levels approaching baseline in the week of July 26, 2020 (median=84.8%, IQR=78.1%, 90.0%), and then decreased slowly through the fall and rapidly in December 2020. Manitoba was unique in Canada, with mobility levels dropping comparatively more during the fall of 2020. Park mobility in all provinces peaked over the summer months (Appendix 1).

**Figure 1.**
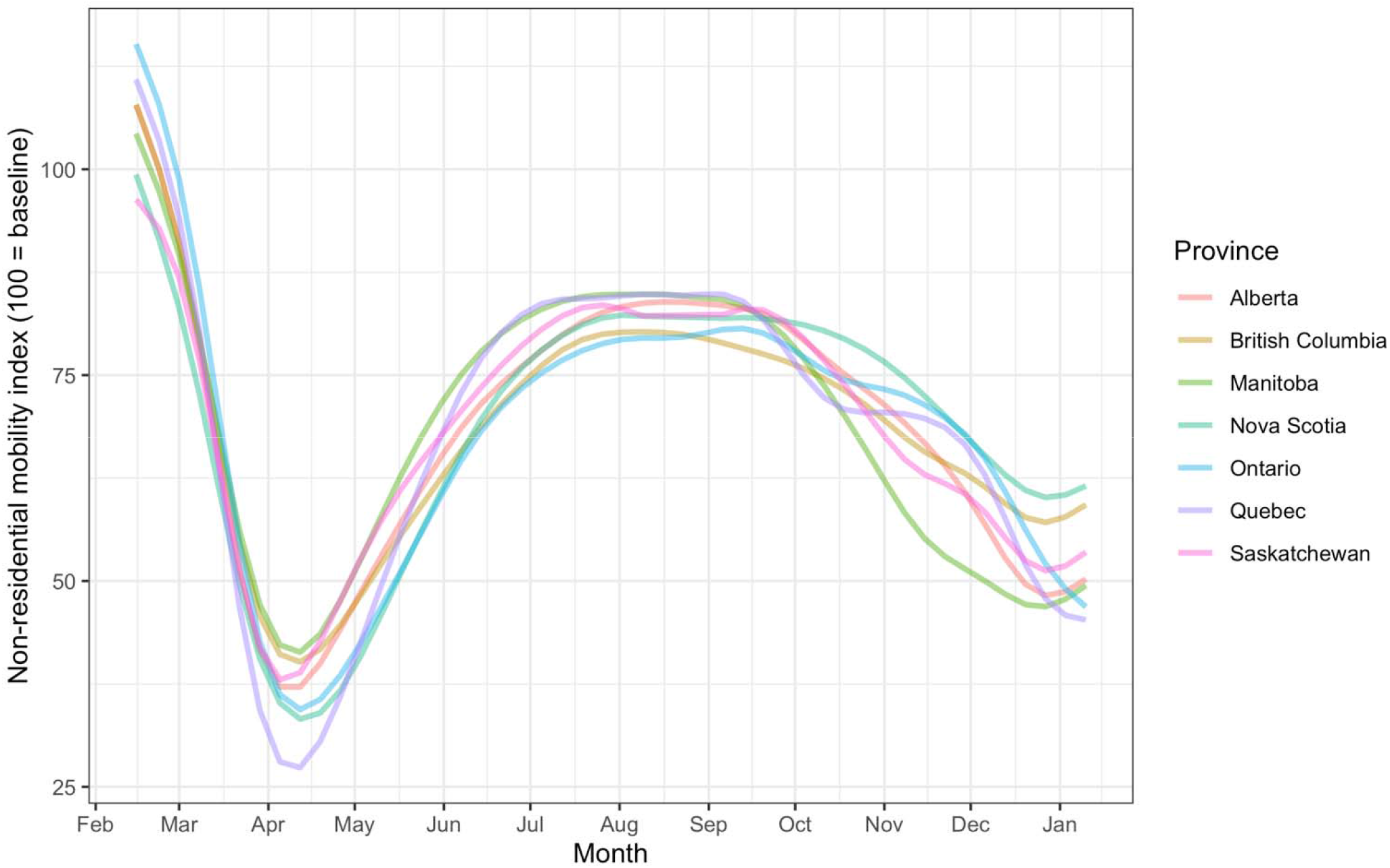
Non-residential mobility across 7 Canadian provinces, February 5, 2020 to January 16, 2021. The non-residential mobility index is a measure of the average amount of time spent outside of home, based on smartphone mobility data (the index is scaled so that levels in the baseline period from Jan 3–Feb 6, 2020 represent 100).

### Mobility and COVID-19 case growth

Before adjustments for province and seasonality (Model 1), each 5% increase in non-residential mobility was associated with a 5% acceleration in COVID-19 growth (GRR = 1.05, 95%CI: 1.03, 1.07; Table 2), but the predictive accuracy of the model was low (R^2^=11.5). Adjustment for province and seasonality greatly increased the predictive accuracy (R^2^=38.0%), and lead to increased observed strength of the mobility association (GRR=1.09, 95%CI: 1.07, 1.12). COVID-19 case growth, for a given time of year and level of mobility, was highest in Ontario (GRR=1.10, 95%CI: 0.98, 1.24) and Quebec (GRR=1.08, 95%CI: 0.96, 1.22). Figure 2 depicts the importance of mobility, after accounting for provincial, and seasonal (monthly) variation in COVID-19 growth rates.

**Table 2.**
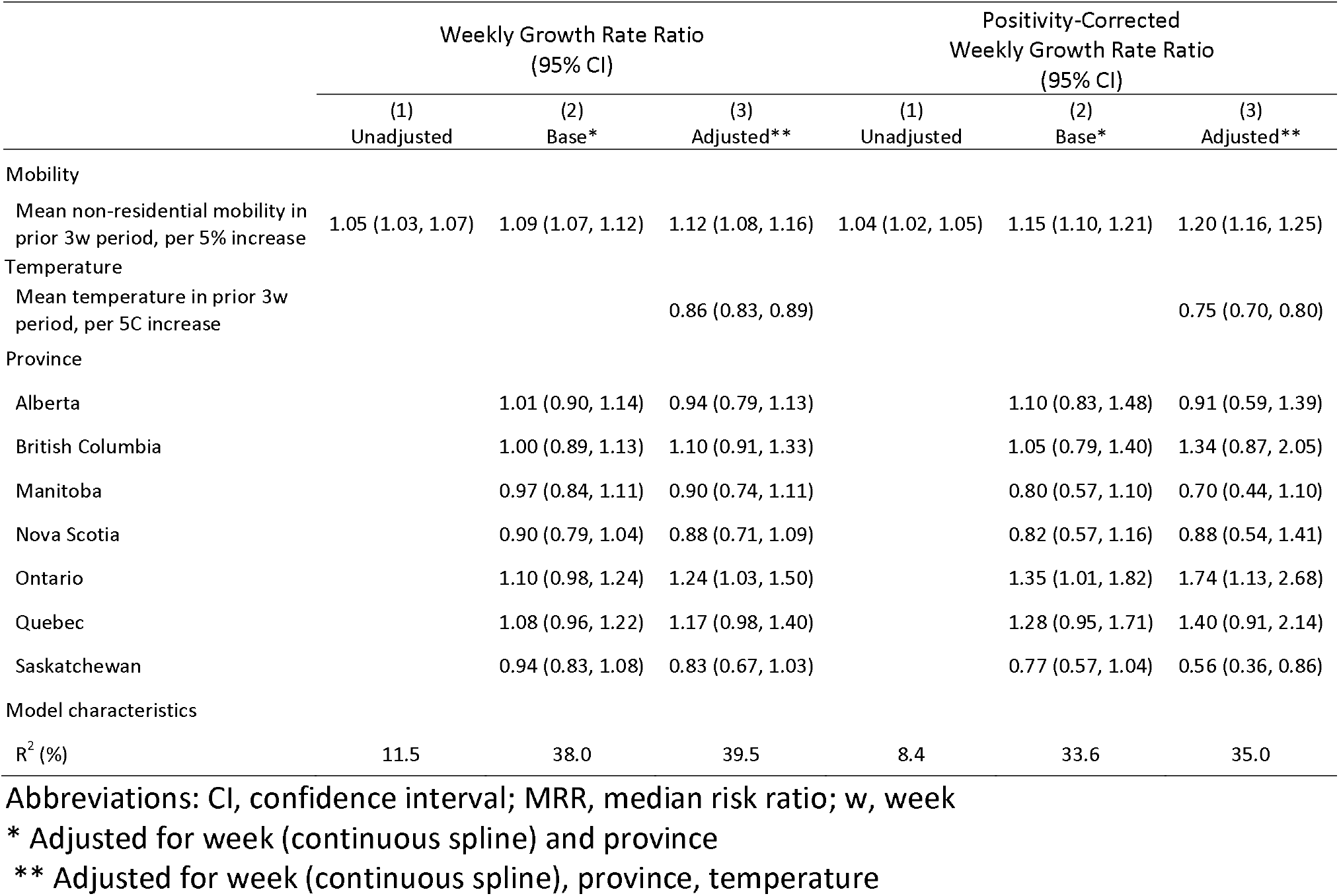
Factors influencing COVID-19 weekly growth rates, and positivity-corrected growth-rates, across 7 Canadian provinces, March 15, 2020 to January 16, 2021.

**Figure 2.**
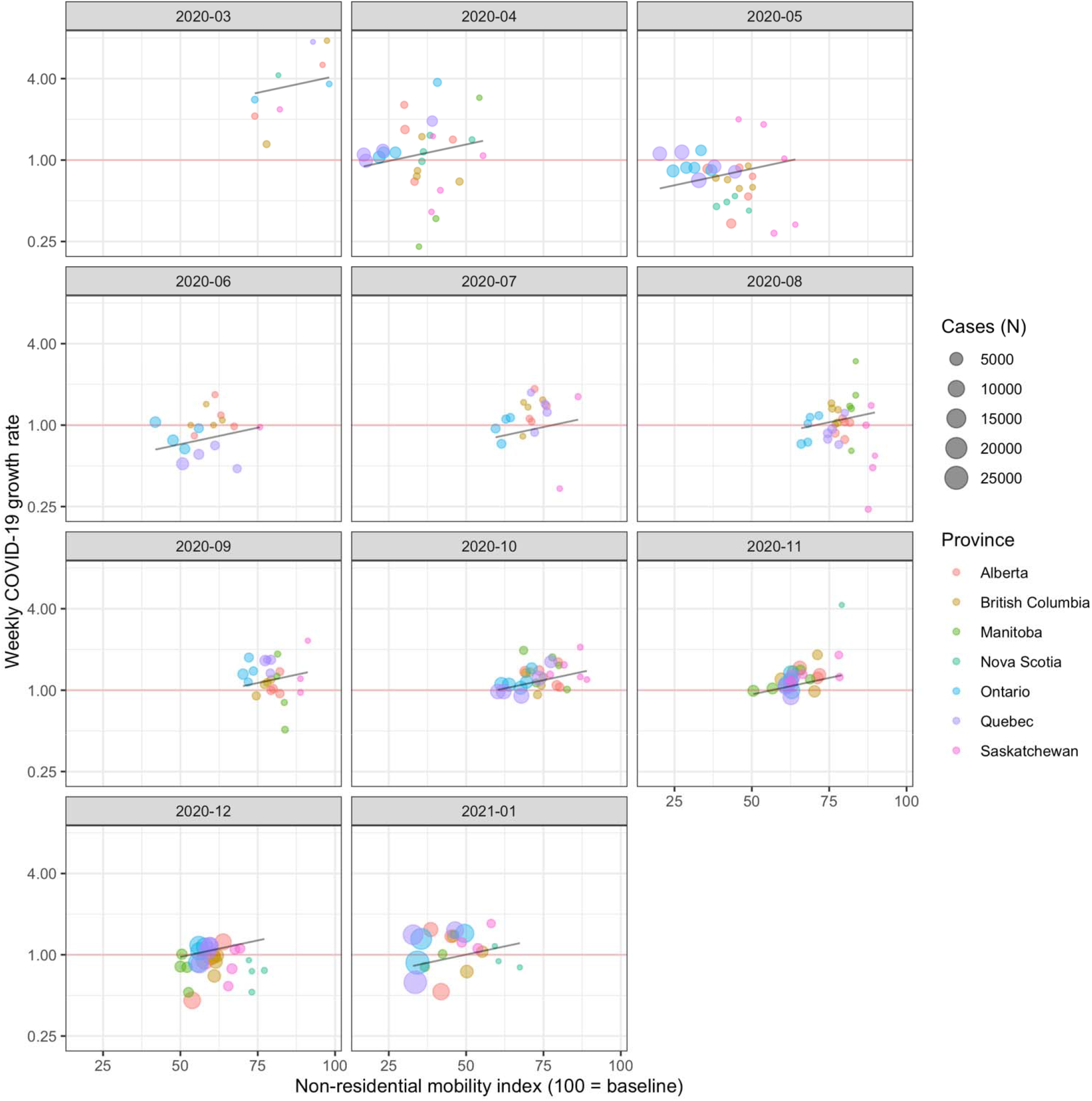
Association between non-residential mobility and COVID-19 case growth across 7 Canadian provinces, March 15, 2020 to January 16, 2021. Weekly COVID-19 growth rate (Y = cases in given week / cases in prior week) is strongly associated with the non-residential mobility in the prior 2-week period (X). The point where the regression line (black) crosses the line representing no COVID-19 case growth (red line) is the average Canadian mobility threshold. The Canadian mobility threshold is lower in spring, fall and winter of 2020, compared to the summer of 2020.

In the model adjusting for temperature effects (Model 3), mobility remained associated with COVID-19 case growth (GRR=1.12, 95%CI: 1.08, 1.16). Increases in mean weekly temperature were significantly associated with decreased COVID-19 growth (GRR=0.86 per 5C increase, 95%CI: 0.83, 0.89). Temperature explained 68.2% of the variation in COVID-19 case growth associated with calendar date (18). In a sensitivity analysis, we used park mobility, instead of temperature in model 3; each 50% increase in park mobility was associated with a 13% reduction in incidence (GRR=0.87, 95%CI: 0.77, 0.97). Analyses that were based on the corrected weekly case growth measure yielded associations that were stronger than in the case growth analysis. Namely, in the base model, each 5% increase in non-residential mobility was associated with a 15% acceleration in corrected COVID-19 growth (95%CI: 1.10, 1.21).

### Mobility threshold

The base growth rate model was used to measure the mobility threshold. The mobility threshold varied markedly through the pandemic period (Figure 3) and was highest in the week of August 2^nd^ (median=72.6, IQR: 70.1, 76.1), and dropped throughout the fall and winter of 2020 to reach low levels in January 2021 (median=52.7, IQR: 49.9, 56.2). Variations across provinces in the estimated mobility threshold were apparent: in the week of January 10, 2021, the lowest mobility thresholds were observed in Ontario (45.4) and Quebec (46.9).

**Figure 3.**
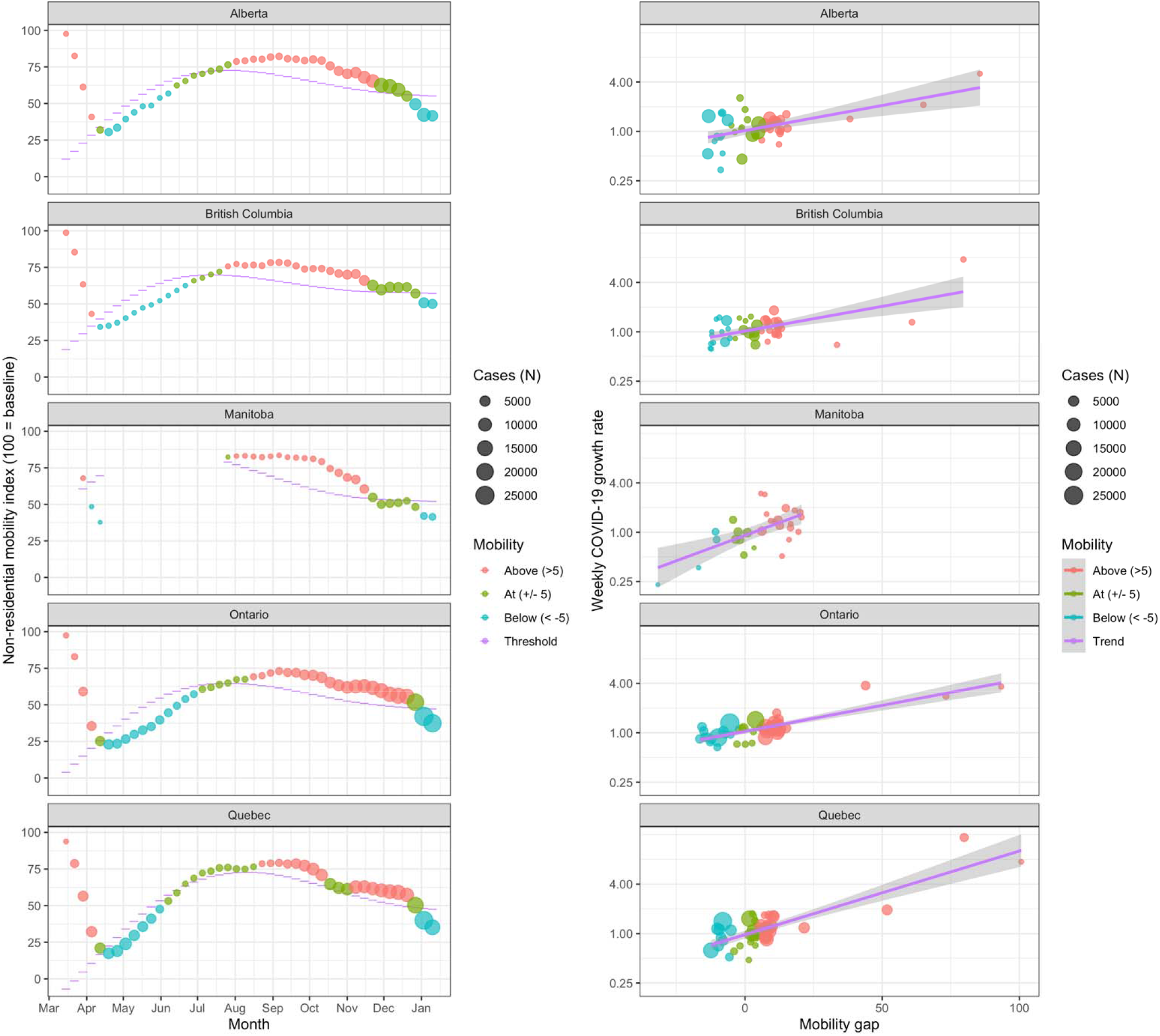
Left panel: Variation in mobility (circles) and the estimated mobility threshold (purple dash), for 5 Canadian provinces with the most cases, March 15, 2020 to January 16, 2021. Right panel: Association between mobility gap and COVID-19 growth rate. The mobility threshold is the estimated level of mobility needed to control COVID-19 case growth. This threshold is highest in summer and in less populated provinces. When mobility decreased below the mobility threshold (blue dots), weekly COVID-19 cases counts decreased. In November 2020, Manitoba was the only province that implemented a lockdown that had successfully crossed the mobility threshold and has led to reductions in COVID-19 case growth. Other provinces waited until December 2020.

As a sensitivity analysis to better understand the impact of the United Kingdom Variant of Concern (VOC) on mobility thresholds, we estimated the reduction in mobility to control a variant with 1.4-fold higher reproduction number (19). Using the base growth rate model for the main outcome, and an estimated generation interval of 5.0 days (20,21), we estimated that an additional 6.8-point (95%CI: 5.2, 8.7) reduction in mobility would be required to control transmission when the VOC prevalence increases to 25%, and an additional 20.5-point (95%CI: 15.6, 26.1) reduction would be required when the VOC prevalence reaches 75%. The adjusted model, which had a larger effect size for mobility, yielded a smaller estimated reduction in order to control COVID-19 case growth (25% VOC: 5.2, 95%CI: 4.0, 7.6; 75% VOC: 15.6: 11.9, 22.9).

### Mobility gap

The mobility gap in Canadian provinces passed through distinct phases (Figure 3) over the course of the pandemic. At the onset of the pandemic in March 2020, non-residential mobility was in excess of the mobility threshold. Strict lockdown measures led to rapid declines in mobility and control of COVID-19 case growth (April-May). Easing of lockdown measures in the late spring of 2020 coincided with lower mobility thresholds, but mobility soon grew in excess of levels needed to control COVID-19 (June-August).

Mobility thresholds decreased throughout the fall and mobility remained above the threshold, coinciding with surging case counts (September-November). In November 2020, Manitoba markedly reduced mobility to levels below the mobility threshold. In January 2021, Quebec and Ontario were the last provinces to achieve mobility reductions that put them below the mobility threshold.

### Validation and Sensitivity Analyses

In a sensitivity analysis to ascertain the robustness of the mobility threshold, all 3 alternative mobility threshold measures were highly concordant with the base measure (Pearson correlation ≥0.94, Appendix 2). As a statistical validation, we verified that the mobility gap was associated with case growth. Indeed, across the seven provinces, the mobility gap was associated with COVID-19 incidence (median R^2^ = 0.45), and showed good calibration, wherein a mobility gap of 0 closely corresponded to no case growth (Figure 3).

## Discussion

In this study we examined predictors of week-over-week COVID-19 case growth across Canadian provinces. Our data suggest that reductions in mobility strongly predict future control of COVID-19 growth in the subsequent 3-week period, and that more substantial reductions in mobility were required to suppress transmission of COVID-19 through fall 2020. In this manuscript, we developed novel measures of the estimated mobility level required to achieve COVID-19 control in seven Canadian provinces (the mobility *threshold*), and the estimated mobility reduction required to control COVID-19 case growth (the mobility *gap*).

This study builds on work showing strong associations between physical distancing measures and COVID-19 incidence. Studies using smartphone mobility measures demonstrate that changes in mobility specifically predict COVID-19 incidence in the subsequent 1-3 weeks (11). More detailed mobility data also suggested that dine-in restaurants, take-out services (likely representing risk for workers more than customers), gyms, and cafes are particularly important drivers of COVID-19 incidence in the United States (22). Our work shows that required mobility reductions are seasonally dependent: only small reductions were required to control COVID-19 in the summer of 2020 but larger mobility reductions have been needed since the fall.

As with several respiratory pathogens (23,24), we observed substantial seasonal variation in baseline risk of COVID-19. Substantial controversy remains as to the underlying drivers of wintertime seasonality; hypotheses for these drivers include human behavioural factors, increased virus survival due to climatic conditions (in particular, decreased absolute humidity) (25), and immune system related factors (23). Using real human behavioural data, we found that weather as well as increased time spent in outdoor settings were important drivers of decreased COVID-19 risk in the summertime, explaining over 50% of seasonal variation risk of COVID-19 (26).

Our work suggests that if governments and public health agencies wish to supress community transmission of COVID-19 in winter months, stringent NPIs may be necessary. Manitoba, which lowered mobility sufficiently to achieve control of COVID-19 in the fall, did so by moving the entire province into the most stringent lockdown level on November 12, 2020. Measures included restricting private gatherings to 5 persons, and closing both non-essential businesses and in-restaurant dining (27), and increased enforcement (almost 1 million dollars in fines given out by early January 2021) (28,29).

Our study has some limitations. First, our study did not examine granular patterns of mobility within provinces, limiting potential insights into the effectiveness of the regional approaches pursued in some provinces. Second, our study used comparative measures of mobility, relative to levels in January 2020, adding complexity to interprovincial comparisons. Third, this study lacked data on COVID-19 vaccination, first administered in Canada on December 14, 2020 and remaining below herd immunity levels in January 2021. As vaccination rates increase, these can be embedded into the model as an increasing mobility threshold. Meanwhile, other factors, such as the rapidly spreading variant arising from the United Kingdom and South Africa (30), may lead to a decreasing mobility threshold needed to control COVID-19.

This study demonstrates that mobility strongly predicts COVID-19 case growth up to 3-weeks in the future, and that stringent measures will continue to be necessary through the winter 2021 months in Canada. The mobility threshold and mobility gap concepts developed here can be used by public health officials and governments to estimate the level of restrictions needed to control COVID-19 and guide, in real-time, the implementation and intensity of NPIs to control COVID-19.

## Data Availability

All data used in this work are publicly available (Google mobility reports, opencovid.ca, R weathercan library)

https://www.google.com/covid19/mobility/

https://opencovid.ca/

## Appendices

Appendix 1. Sensitivity analysis: alternative mobility threshold models

We compared the mobility threshold based on the case growth model incorporating temperature to the base mobility threshold model. Measurement of the mobility threshold based on the model 3 yielded results that were largely identical (Pearson correlation = 0.96) to the primary threshold model. We did a similar sensitivity analysis, based on models 2 and 3 for the corrected case growth outcome. Again, measurement of the mobility threshold based on the corrected weekly case growth outcome was nearly identical to the base model (model 2: Pearson correlation = 0.97; model 3: Pearson correlation = 0.94).

**Appendix 2.**
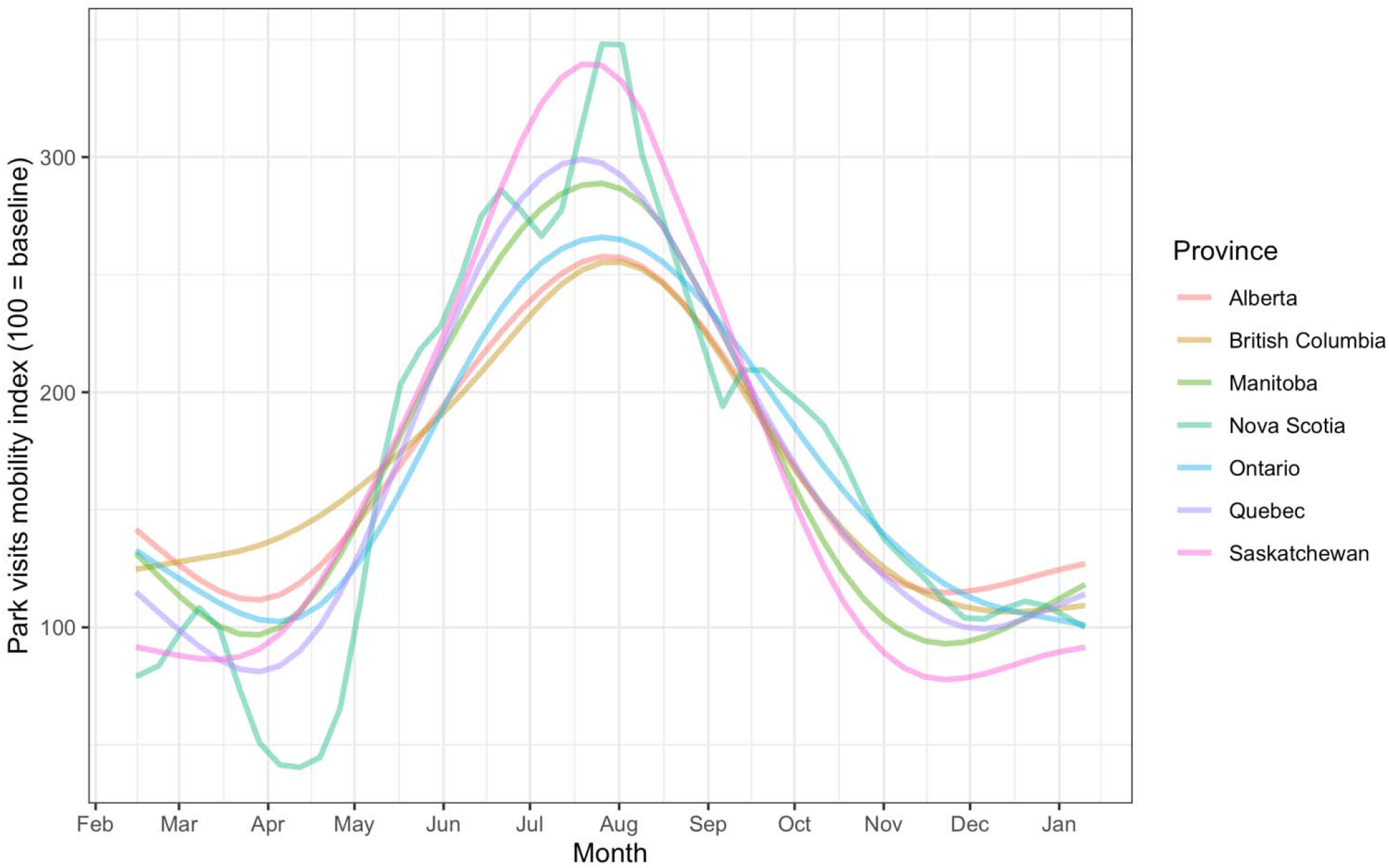
Park visits mobility across 7 Canadian provinces, February 5, 2020 to January 16, 2021. The park visits mobility index is a measure of the frequency of visits to locations classified as parks across the entire provincial population, based on smartphone mobility data (the index is scaled so that levels in the baseline period from Jan 3–Feb 6, 2020 represent 100).

## Notes

### Competing Interest Statement

The authors have declared no competing interest.

### Funding Statement

No external funding was received for this work.

### Author Declarations

Projects based on publicly available data do not require Public Health Ontario or University of Toronto IRB review.

